# Quantification of neuromotor control in *STXBP1*-Related Disorders with wearable sensors

**DOI:** 10.1101/2025.09.20.25336193

**Authors:** Julie M. Orlando, Bintou Bane, Torrey Chisari, Jan H. Magielski, Samuel R. Pierce, Kristin Cunningham, Charlene Woo, Sarah Tefft, Joeylynn Nolan, Sarah Ruggiero, Michael J. Boland, Jillian L. McKee, Ingo Helbig, Laura A. Prosser

**Affiliations:** Division of Rehabilitation Medicine, Children’s Hospital of Philadelphia, Philadelphia, PA, 19104, USA; Center for Epilepsy and Neurodevelopmental Disorders (ENDD), University of Pennsylvania Perelman School of Medicine and Children’s Hospital of Philadelphia, Philadelphia, PA, USA; Division of Neurology, Children’s Hospital of Philadelphia, Philadelphia, PA, 19104, USA; The Epilepsy NeuroGenetics Initiative (ENGIN), Children’s Hospital of Philadelphia, Philadelphia, PA, 19104, USA; Department of Biomedical and Health Informatics (DBHi), Children’s Hospital of Philadelphia, Philadelphia, PA, 19146, USA; Department of Neurology, University of Pennsylvania Perelman School of Medicine, Philadelphia, PA, 19104, USA; Department of Pediatrics, University of Pennsylvania Perelman School of Medicine, Philadelphia, PA, 19104, USA

**Keywords:** epilepsy, genetics, developmental and epileptic encephalopathy, *STXBP1*, movement disorders, neuromotor control

## Abstract

*STXBP1*-Related Disorders (*STXBP1*-RD) are a spectrum of neurodevelopmental conditions that often present with prominent motor impairments which affect fine and gross motor development and are in part attributed to abnormalities of neuromotor control, such as involuntary movement, dystonia, tremor, and ataxia. There is a lack of precise, scalable measurement tools to characterize motor control abnormalities, particularly in children with physical and intellectual disability who cannot complete traditional testing protocols, which has contributed to an incomplete understanding of movement disorders within the broader disease trajectory of *STXBP1*-RD. Emerging wearable sensor technology has the potential to meet this need.

Here, we quantified the prevalence and clinical features of tremor and other motor control impairments in 31 individuals with *STXBP1*-RD, 64.5% with tremor, compared to 19 typically developing controls during a simple reaching task using inertial sensors on the upper arms and forearms. We then evaluated the clinical relevance of the sensor-derived metrics of motor control in the context of traditional developmental clinical assessments and additional data sources, including previously published retrospective studies, aggregated EMR data, and a prospective natural history study.

Individuals with *STXBP1*-RD demonstrated slower reaching time, decreased arm acceleration intensity, less smooth reaching, and tremor frequency characteristics consistent with an unstable or irregular rhythmic movement pattern. Combining features from all four upper limb sensors predicted tremor with the highest accuracy (AUC = 0.87) over combinations with fewer sensors, and multiple motor control metrics were significantly related to clinical assessment scores, including those from the Bayley Scales of Infant and Toddler Development, Peabody Developmental Motor Scales and the Gross Motor Function Measure-66. We also found that more than 65% of individuals with *STXBP1*-RD present with tremor in the natural history study cohort. This prevalence exceeds by fivefold what has been reported in other genetic neurodevelopmental disorders, suggesting that tremor may be heavily underreported in historical, retrospective datasets.

Our study provides a novel understanding of the motor control features in one of the most common genetic neurodevelopmental disorders and establishes a robust paradigm to quantify neuromotor control in individuals with developmental differences. Our observations demonstrate strong relations between motor control measures and developmental function in this population. With a range of personalized therapies currently in development for *STXBP1*-related disorders, assessment of neuromotor control and tremor in this cohort could serve as biomarkers for future clinical trials.

## Introduction

Individuals with developmental and epileptic encephalopathies (DEEs) present with a broad spectrum of genetic and clinical features, involving over 800 genetic etiologies.^1^ Movement disorders are often part of the heterogenous phenotypic presentation found in individuals with DEEs, and there has been a recent emphasis on identifying and characterizing these movement disorders. Overall, the presence of movement disorders varies by diagnosis with broader terms, such as abnormality of movement being reported more frequently than more specific terms such as tremor.^2,3^ The most commonly observed movement disorders within DEEs are classified as hyperkinesias and include repetitive stereotypies, dystonia, chorea, myoclonus, ataxia, and tremor.^4–6^ Strikingly, about half of individuals have more than one movement disorder.^6^ This growing focus on classifying movement disorders may be attributed to emerging advancements in therapeutics necessitating a need to establish disease-specific biomarkers and define meaningful subgroups within DEEs clinical populations to evaluate therapeutic efficacy.^7,8^

*STXBP1*-Related Disorders (*STXBP1*-RD) are a spectrum of neurodevelopmental conditions caused by pathogenic mutations in the *STXBP1* gene. First described in 2008 in a group of individuals with Ohtahara Syndrome and neonatal epilepsy,^9^ *STXBP1*-RD are now known to be one of the most prevalent monogenic causes of neurodevelopmental disorders and epilepsy. Individuals often present with seizures, developmental delays, intellectual disability, and hypotonia. Individuals with *STXBP1*-RD commonly display impairments in neuromotor control or specified movement disorders, including involuntary movements, stereotypies, dystonia, ataxia, and tremor.^10^ ^5,11–16^ These impairments have been described through case studies and retrospective studies as well as reported in interviews with families of individuals with *STXBP1*-RD and healthcare providers.^10,11,13,17^

Few studies have sought to quantify or characterize the features of neuromotor impairments or movement disorders in individuals with *STXBP1*-RD. Loussouarn et al., conducted neurophysiological evaluations on six individuals with *STXBP1*-RD with tremor.^18^ Participants presented with distal or proximo-distal rhythmic myoclonus when holding their hands outstretched or performing an action (e.g., drinking), and EMG features included regular or irregular bursts with a large duration range and frequencies of 4-10 Hz. The authors concluded that observed tremor was most likely a subcortical myoclonus based on the EMG patterns. The authors also suggested that ataxia, which is often described clinically may also be myoclonus, though this hypothesis has not been confirmed.^18^ The ambiguity about the movement impairments that characterize *STXBP1*-RD illustrates the need to develop tools to identify and objectively characterize these unique movement patterns.

Objective characterization has been limited by a lack of consensus on how to describe the clinically observed movement disorders, specifically, ataxia and tremor often observed in individuals with *STXBP1*-RD. Current clinical descriptions include relative location and duration of the tremor such as greater prominence in the upper extremities than the lower extremities and the duration of the tremor, such as present throughout a clinical examination. Clinicians may also describe the potential impact of tremor on daily activities but have not been able to quantify tremor or motor control impairments in a measurable way. Importantly, these descriptions are relatively subjective, without established validity and reliability and with wide variability in descriptive approach among clinicians.

While standardized rating scales exist to assess tremor and ataxia, these scales require following complex multi-step instructions to perform coordination tasks are therefore not feasible for individuals with *STXBP1*-RD due to their language and cognitive impairments.^19,20^ Even modified rating scales such as the Scale for Assessment and Rating of Ataxia in Toddlers include items that are not feasible for the *STXBP1*-RD patient population due to their physical and intellectual disabilities.^21^ This lack of precise measurement has contributed to an incomplete understanding of movement disorders within the broader disease trajectory of *STXBP1*-RD.

Wearable sensors have accurately quantified tremor in several clinical populations with tremor, ataxia, and stereotypies, underscoring the potential of unobtrusive, objective tools to fill these critical gaps.^22^ Wearable sensors worn on the upper limbs have been able to distinguish between parkinsonism and essential tremor when participants maintained resting posture, held their arms outstretched, or performed specific tasks including upper extremity movement patterns and drawing a spiral.^23,24^ Sensor metrics related to posture, locomotion, and upper limb activity collected during daily activities were predictive of clinical measures and biological markers of disease severity in individuals with Friedrich’s Ataxia.^25^ Similarly, gait ataxia and upper limb motor function have been evaluated using wearable sensors in individuals with cerebellar ataxia.^26^ Several studies have used wearables to monitor and classify repetitive stereotypies such as hand-flapping or hand-clenching in individuals with autism spectrum disorders (ASD)^27^ or developmental disabilities,^28^ yet this work is still emerging and studies have been limited to small sample sizes establishing validity. The use of wearable sensors to identify movement disorders or classify movement patterns is promising and continuously growing.^29,30^ Importantly, wearable sensors have been well-tolerated in pediatric clinical populations with ASD^19^, cerebral palsy^31^ and muscular dystrophy.^32^ Notably, wearable sensor metrics were more responsive to change than clinical rating scales in detecting medial-lateral gait amplitude over a 1.5-year interval in adults with spinocerebellar ataxia.^33^ This supports the potential to use metrics derived from wearable sensor data to develop valid and objective measures to identify tremor in individuals with *STXBP1*-RD.

Here, we aimed to quantify neuromotor control, including tremor in individuals with *STXBP1*-RD compared to individuals with typical development (TD) during a structured reaching task. We assessed the clinical relevance of sensor-derived motor control metrics by examining their relationships with standardized developmental assessments and contextualizing tremor prevalence using previously published retrospective studies, aggregated electronic medical record data, and a prospective natural history study.

## Materials and methods

### Prospective Cohort Participants

All Participants were enrolled in the multi-site STARR Natural History Study for *STXBP1*-RD (Clinical Trials Identifier: NCT06555965). All participants were seen at a single site, the Children’s Hospital of Philadelphia. A subset of STARR participants also enrolled in the current study involving wearable sensor–based movement assessment. Recruitment for this study occurred during scheduled STARR visits and informed consent was obtained from parents or legal guardians prior to participation. The inclusion criteria were: confirmed diagnosis of *STXBP1*-RD through genetic testing and verified by an expert genetic counselor or neurologist. Participants with TD were recruited through convenience sampling, and the inclusion criterion was age-appropriate milestone achievement with no developmental concerns as reported by the parent or caregiver. Exclusion criteria for both groups were: (1) history of prematurity (defined as gestational age <35 weeks), interventricular hemorrhage, structural brain deficit, or congenital heart disease; and (2) behavioral concerns (e.g., throwing, banging) with a high likelihood of damaging the sensors, as determined by a clinician, (3) inability to reach at least three times with either upper extremity during the reaching assessment.

### Assessment Procedures

Wearable sensors were about the size and weight of a smartwatch and contained a tri-axial accelerometer, gyroscope, and magnetometer (Opal, Clario). Sensors were donned on the participant’s upper arms and forearms and secured with Velcro straps (**Figure 1**). Data were collected asynchronously with a sampling rate of 20 Hz, stored on the devices, and then uploaded onto a secure server for signal processing. Participants were seated in a supportive chair, their own personal mobility device or if preferred by the participant, on a floor mat without additional support from another person. Participants were then encouraged to reach to a target that was positioned to elicit 90 degrees of shoulder flexion (i.e., toy at shoulder-height and approximately 90% of their arm length). A selection of hand-held toys was used to elicit reaching; however, if the participant did not reach, another preferred object was used, such as a snack, water bottle, or a favorite toy. Examiners attempted to elicit a minimum of three reaches with each upper extremity using tactile and verbal cues. The data collection procedures were video recorded.

**Figure 1.**
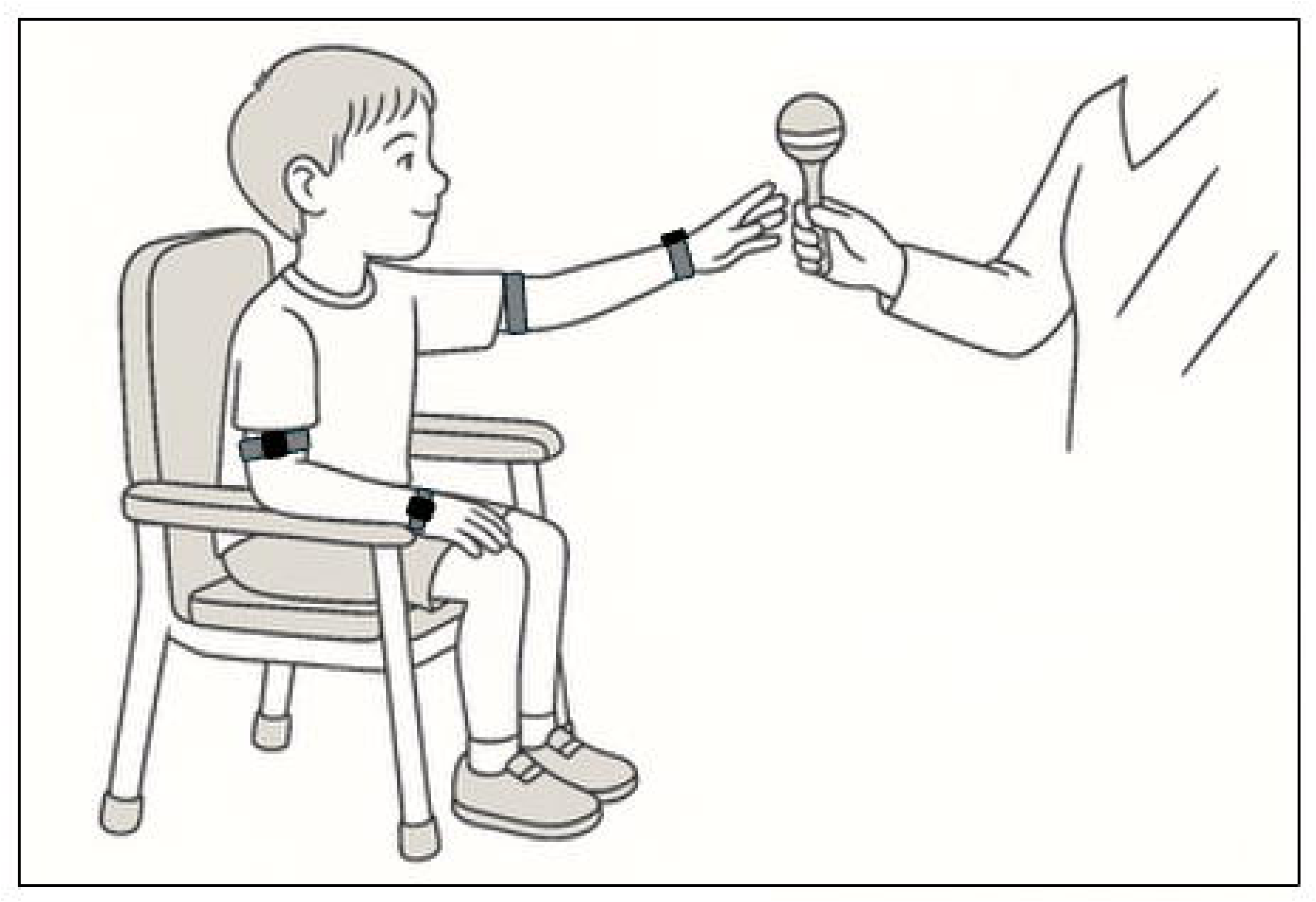
Structured Reaching Task Schematic. The participant is seated in a supportive chair with sensors donned at his forearms and upper arms bilaterally and is reaching to a target that that is positioned to elicit a reach with 90 degrees of shoulder flexion.

### Identifying Reaching Intervals through Behavioral Coding

Behavioral coding to identify reaching intervals was completed using Datavyu behavioral coding software.^34^ A reaching interval was operationally defined as, an individual moving their hand forward from a resting position towards the target object positioned at shoulder height within arm’s length and the reach was considered complete when the hand contacted the object. If the participant demonstrated a clear change in movement direction away from the toy without subsequent correction toward the target, the reaching interval was considered incomplete and was not evaluated. The position of the participant was also noted as either seated in a chair or seated on the floor. If the reach occurred in another position (such as on a caregiver’s lap or standing), the reach was not evaluated.

### Motor Coordination Metrics Derived from Wearable Sensor Data During Reaching Intervals

For each participant, raw tri-axial acceleration and angular velocity data were extracted from the four wearable sensors (dominant upper arm, dominant forearm, non-dominant upper arm, and non-dominant forearm) during completed reaching intervals. The raw acceleration and angular velocity signals were not filtered due to the relatively low sampling rate and the small number of datapoints within each reach bout.

Median reach duration was computed separately for the dominant and non-dominant arms. The dominant arm was operationally defined as the limb with the faster median reach duration across all recorded trials. For each arm, the three reaches with durations closest to the median were selected for further analysis. Raw sensor data from these three reaches were concatenated for each sensor, resulting in one continuous time series per sensor per participant. Sensor-based metrics were then computed on these concatenated datasets to characterize the movement behavior of each limb segment.

Motor coordination metrics were chosen to capture different aspects of movement, including duration, intensity, smoothness, and tremor characteristics. **Table 1** describes the constructs, definition, interpretation, and the method of calculation for each metric. All Metrics were robust to short-duration time series signals, with a minimum threshold of approximately 30 data points for reliable computation.

**Table 1.**
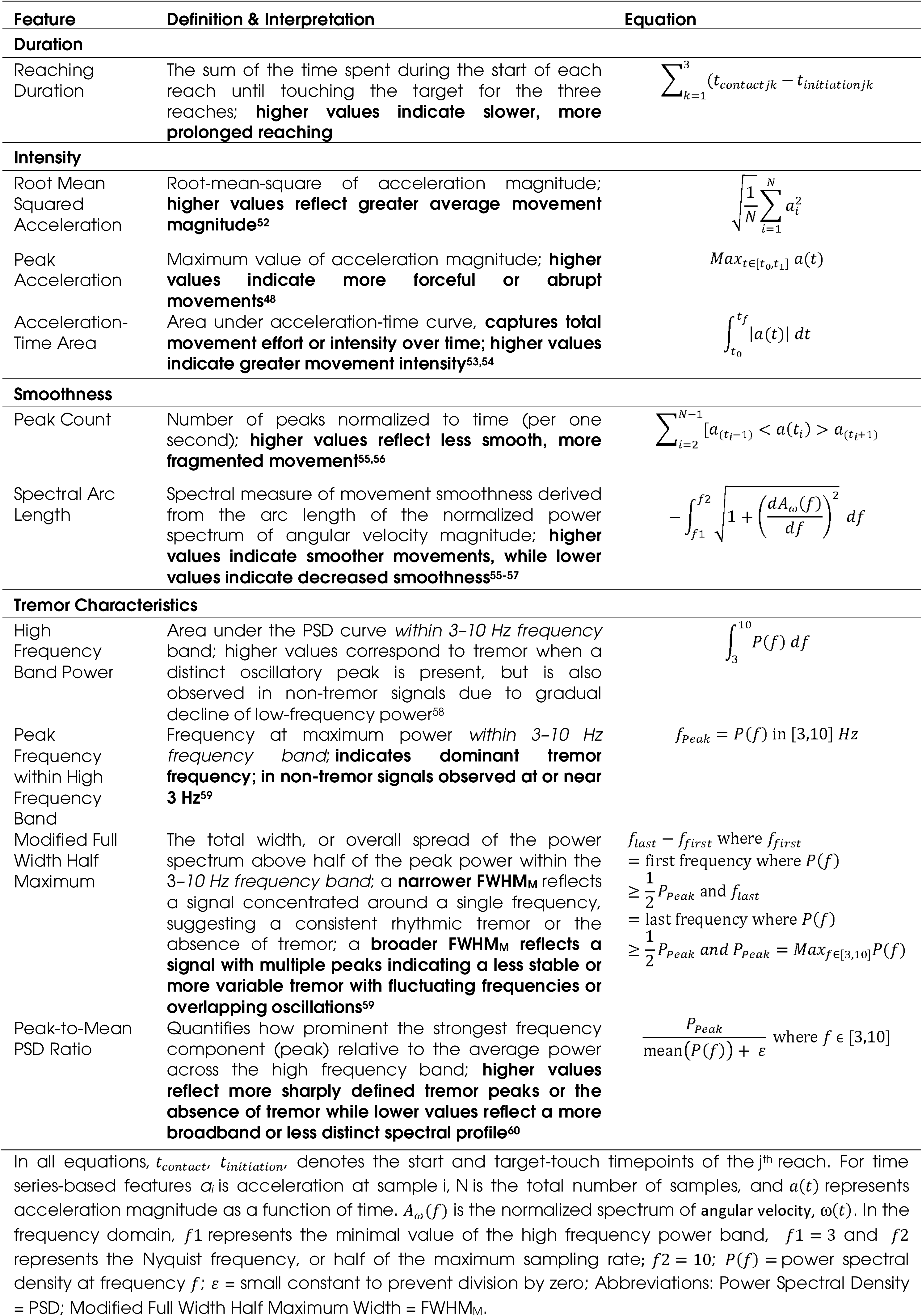
Sensor-Derived Metrics Summary.

### STXBP1-Clinical Severity Assessment

As part of the STARR study visit at this site, the presence of tremor was evaluated using a standardized rating scale. The *STXBP1*-Clinical Severity Assessment (S-CSA) is a clinician-reported outcome measure designed to evaluate motor, communication, and vision domains,^35,36^ which is derived from the CDKL5 Clinical Severity Assessment.^37^ It is administered by pediatric neurology experts (MD or APP) who have been trained in its use. For the current study, we utilized the subsection relating to tremor within the S-CSA to obtain standardized assessment of the movement abnormalities. The S-CSA includes a rating scale that characterizes tremor based on its distribution, duration, and functional impact. For the present study, tremor distribution classifying tremor as absent, focal, segmental, or generalized was used.

### Developmental Assessments

All developmental assessments were conducted by assessors who completed assessment specific training as part of the STARR study. The Bayley Scales of Infant and Toddler Development, 4^th^ Edition (Bayley-4) was used to evaluate gross motor, fine motor, cognition, expressive language and receptive language development with normative-reference age-equivalent scores.^38^ Gross motor development was also evaluated with the criterion-referenced Gross Motor Function Measure-66 (GMFM-66) total score, administered by a trained physical therapist.^39^ The Peabody Developmental Motor Scales, 3^rd^ Edition (PDMS-3), hand function and eye-hand-coordination subscales were administered by an occupational therapist and converted to normative-reference age equivalent scores.^40^ All assessments were conducted on the same day as the wearable sensor data collection.

### Movement Disorder Population Prevalence

To identify the prevalence of movement disorders in individuals with *STXBP1*-RD compared to other NDDs, we compared the frequencies of established terminology across different datasets. Retrospective cohort data were obtained previously published literature^10^ and from the Citizen Health^41^ dataset, a patient-reported data platform that aggregates real-world clinical information from individuals with neurodevelopmental disorders (NDDs), including developmental and epileptic encephalopathies (DEEs) of presumed genetic origin. Data extraction focused on standardized Human Phenotype Ontology (HPO)^42^ terms related to movement disorders: abnormality of movement (HP:0100022), tremor (HP:0001337), ataxia (HP:0001251), and myoclonus (HP:0001336). Frequencies of these terms were reviewed across several genetic NDD diagnoses, including *SCN8A*-Related Disorders, *SYNGAP1*-RD, and *STXBP1*-RD. The prevalence of tremor in the STARR Natural History Study cohort from our site, The Children’s Hospital of Philadelphia, reported via the S-CSA as described above, was also calculated.

### Statistical Analysis

All sensor metric computations were performed using a custom MATLAB script. Statistical analyses were then conducted using R Statistical Framework. Computer code for all analysis is available at github.com/BLINDED.

Preliminary data screening included assessment of normality for all continuous variables using the Shapiro-Wilk test within diagnostic groups (*STXBP1*-RD, TD). Descriptive statistics were reported as means and standard deviations for normally distributed variables, and medians with interquartile ranges for non-normally distributed variables. To evaluate differences in reach durations between dominant and non-dominant limbs, paired-sample t-tests were conducted for normally distributed difference scores; Wilcoxon rank-sum tests were used when normality assumptions were violated.

For between-group comparisons (*STXBP1*-RD vs. TD) of sensor-derived motor control metrics, independent-samples t-tests were applied to normally distributed metrics, while Wilcoxon rank-sum tests were employed for non-normally distributed metrics. Multiple comparisons were addressed using the Benjamini-Hochberg false discovery rate (FDR) correction (q = 0.05).

For each motor control metric, individual participant values were standardized to the TD reference group by calculating z-scores. Each participant’s score reflects the number of standard deviations their performance deviated from the TD mean. Metrics with zero variance in the TD group were excluded.

Machine learning analyses were performed using a supervised Random Forest classifier to evaluate the predictive value of the sensor-based motor control metrics, as described in **Table 1**, for clinically rated tremor presence. We evaluated nine models based on different sensor configurations –Bilateral upper arm and forearm sensors, non-dominant upper arm and forearm sensors, dominant upper arm and forearm sensors, bilateral upper arm sensors, bilateral forearm sensors, and each sensor individually (non-dominant upper arm, non-dominant forearm, dominant upper arm, and dominant forearm sensor). Receiver Operating Characteristic (ROC) curves were generated to assess model performance, and area under the curve (AUC) values were used as the primary performance metric. Feature importance was calculated for each trained random forest model using the varImp() function in the caret package. Importance was estimated by measuring how much model accuracy dropped when the values for a given feature were randomly shuffled in the validation data not used to train that part of the model. This process was repeated across all cross-validation folds and repetitions, and scores were averaged to produce an overall importance value for each feature within a model. To assess stability across models, we calculated the mean importance for each feature across the sensor configurations, along with the standard deviation.

Associations between sensor-derived metrics of motor control and standardized developmental assessment scores (GMFM-66 total score, Bayley-4 age equivalents, PDMS-3 age equivalents) were evaluated using Spearman’s rank-order correlations, given that age-equivalent scores are ordinal data and the non-normal distribution of the GMFM-66 total scores and most sensor metrics. Significance thresholds for correlations were set at *P* < 0.05, with multiple comparisons addressed using the Benjamini-Hochberg false discovery rate correction (*q* = 0.05). Finally, the prevalence rates of movement disorders across different datasets are presented descriptively.

## Results

### Sensor-Based Assessment was Well-Tolerated Across Ages and Developmental Abilities

Thirty-one participants with *STXBP1*-RD (Mean Age: 8.5 years, SD: 6.1, Range: 1-28) and 19 participants with TD (9.2 years, SD: 4.9, Range: 1-17) were enrolled. Of the participants with *STXBP1*-RD, 5 participants (16.1%) were unable to complete at least 3 reaches with either arm and therefore were excluded from this analysis. Two participants with *STXBP1*-RD were able to complete at least three reaches with only their dominant arm; their available data were included. Data collection procedures were completed within approximately 30 minutes, and all participants tolerated the sensors during the assessment. Sensor data were unable to be evaluated for three additional participants with *STXBP1*-RD due to technology failures, such as battery or recording errors, resulting in a total of 23 participants with *STXBP1*-RD with evaluable sensor data.

The participants with *STXBP1*-RD who completed the reaching task with at least one arm had variable developmental abilities, within the functional range expected in this population.^43^ Developmental performance was evaluated by the Bayley-4 and PDMS-3 age-equivalent scores and the GMFM-66 total score, on which a typically developing 5-year-old would be expected to score 100.^39,44^ Developmental performance ranged from 9.2 to 14.1 months (SD: 5.2–9.4) across domains on the PDMS-3 and Bayley-4, with the highest average age-equivalent in the gross motor domain (14.1, SD: 8.1) and the lowest in the eye-hand coordination domain (9.2, SD: 5.2); GMFM-66 mean total score was 53.6 (SD: 11.0) (**Supplemental Material A**).

### Motor Control Metrics Distinguished *STXBP1-*RD from Controls

First, we evaluated if there were differences between the duration of the dominant or non-dominant arm reaches for participants in both the TD and *STXBP1*-RD groups. For the *STXBP1*-RD group, the difference scores (non-dominant minus dominant arm reach duration) were approximately normally distributed (W = 0.952, *P* = 0.527). A paired t-test indicated no significant difference between dominant and non-dominant limb durations, t(15) = −1.87, *P* = 0.121, with a mean difference of −0.82 seconds (95% CI: [−1.87, 0.24]) between arms. For the control group of individuals with TD, the difference scores were also approximately normally distributed (W = 0.926, *P* = 0.168). A paired t-test similarly showed no significant difference between dominant and non-dominant durations (t(17) = 0.29, *P* = 0.774) with a mean difference of 0.05 seconds (95% CI: [−0.33, 0.43]). Together, these findings suggest that there were no significant asymmetries in reach duration for either diagnostic group. Therefore, we compared sensor metrics between groups using reaching data from the dominant arm. An example of motor control metrics from age-matched, 5-year-old participants with *STXBP1*-RD and TD is presented in **Figure 2**.

**Figure 2.**
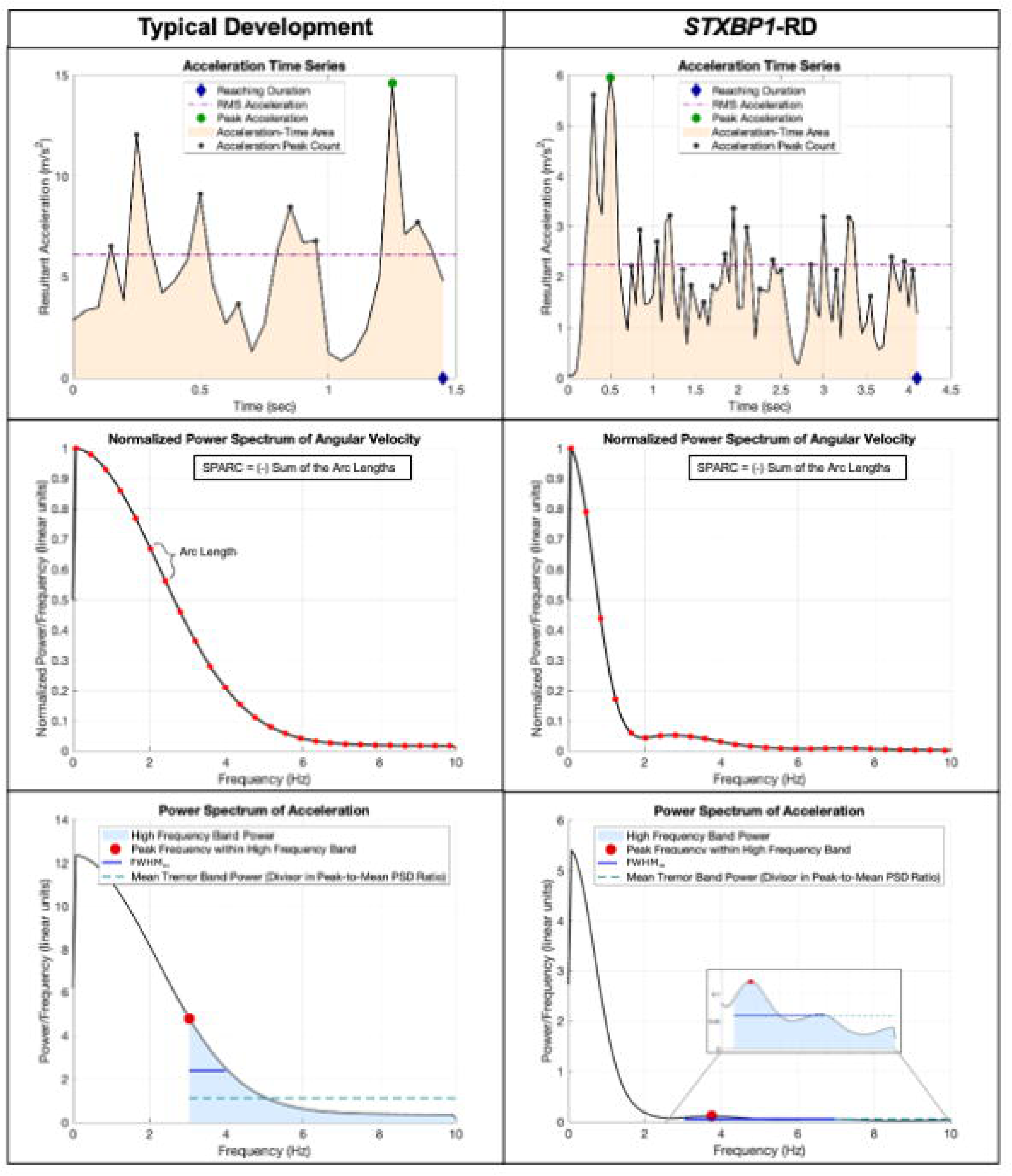
Exemplar time series data for quantification of motor control. (**A**) Acceleration Time Series for a Participant with TD and a participant with *STXBP1*-RD with tremor. TD shows faster reaching duration, similar RMS acceleration, greater peak acceleration, and greater Acceleration Peak Count (normalized by duration, not shown). (**B**) Normalized Power Spectrum of Angular Velocity for the same participants with TD and *STXBP1*-RD with tremor. Examples of the arc length distance on the PSD plot are shown in red. Spectral arc length is the negative sum of the spectral arc lengths and quantifies overall spectral smoothness. (**C**) Power Spectrum of Acceleration for the same participants with TD and *STXBP1*-RD with tremor. **The** normalized power spectrum is in blue with the tremor band (3–10 Hz, light blue) highlighting the area under the PSD curve. Tremor band power is greater for the participant with TD due to greater amplitude at low-frequencies and the gradual decrease across frequencies, compared to the participant with *STXBP1*-RD. The peak acceleration within the high frequency band (red dot) is at a threshold for the high frequency band for the participant with TD. The mean power within the high frequency band (green dashed line) is the divisor in the peak-to-mean PSD ratio and was greater for the participant with TD. The FWHM_M_ was greater for participant with STXBP1-RD, consistent with variable, broadband tremor. Typical Development = TD; *STXBP1*-RD = *STXBP1*-Related Disorders Root Mean Squared = RMS; Power Spectral Density = PSD; Modified Full Width Half Maximum = FWHM_M_.

We assessed the normality of each sensor metric to inform statistical test selection. All metrics were non-normally distributed in each diagnostic group except for reach duration and the time-normalized count of acceleration peaks. We therefore used non-parametric tests to evaluate group differences for non-normally distributed metrics, and parametric tests to evaluate group differences for normally distributed metrics. Reaching duration was significantly slower in the *STXBP1*-RD group (t(29.13) = 6.91, *P* < .001; *STXBP1*-RD: M = 4.08, SD = 1.39; TD: M = 1.92, SD = 0.51). Statistically significant group differences were also observed for all other sensor metrics except for one measure of intensity, the acceleration-time area (V = 756, *P* = 0.765; *STXBP1*-RD: M = 6.6, SD = 3.95; TD: M = 6.81, SD = 3.99) (**Table 2**; **Figure 3**). In contrast, other measures of intensity, peak acceleration and root mean squared (RMS) acceleration, were significantly lower in individuals with *STXBP1*-RD (Peak Acceleration: V = 546, *P* = 0.023; *STXBP1*-RD: M = 5.78, SD = 3.00; TD: M = 8.49, SD = 5.45; RMS Acceleration: V = 350, *P* < .001; *STXBP1*-RD: M = 2.04, SD = 1.14; TD: M = 4.13, SD = 2.82). These findings suggest that while overall movement intensity, as captured by acceleration-time area, may not differ between groups, the maximum instantaneous acceleration and sustained movement intensity are reduced in individuals with *STXBP1*-RD, consistent with hypokinetic movement patterns.

**Figure 3.**
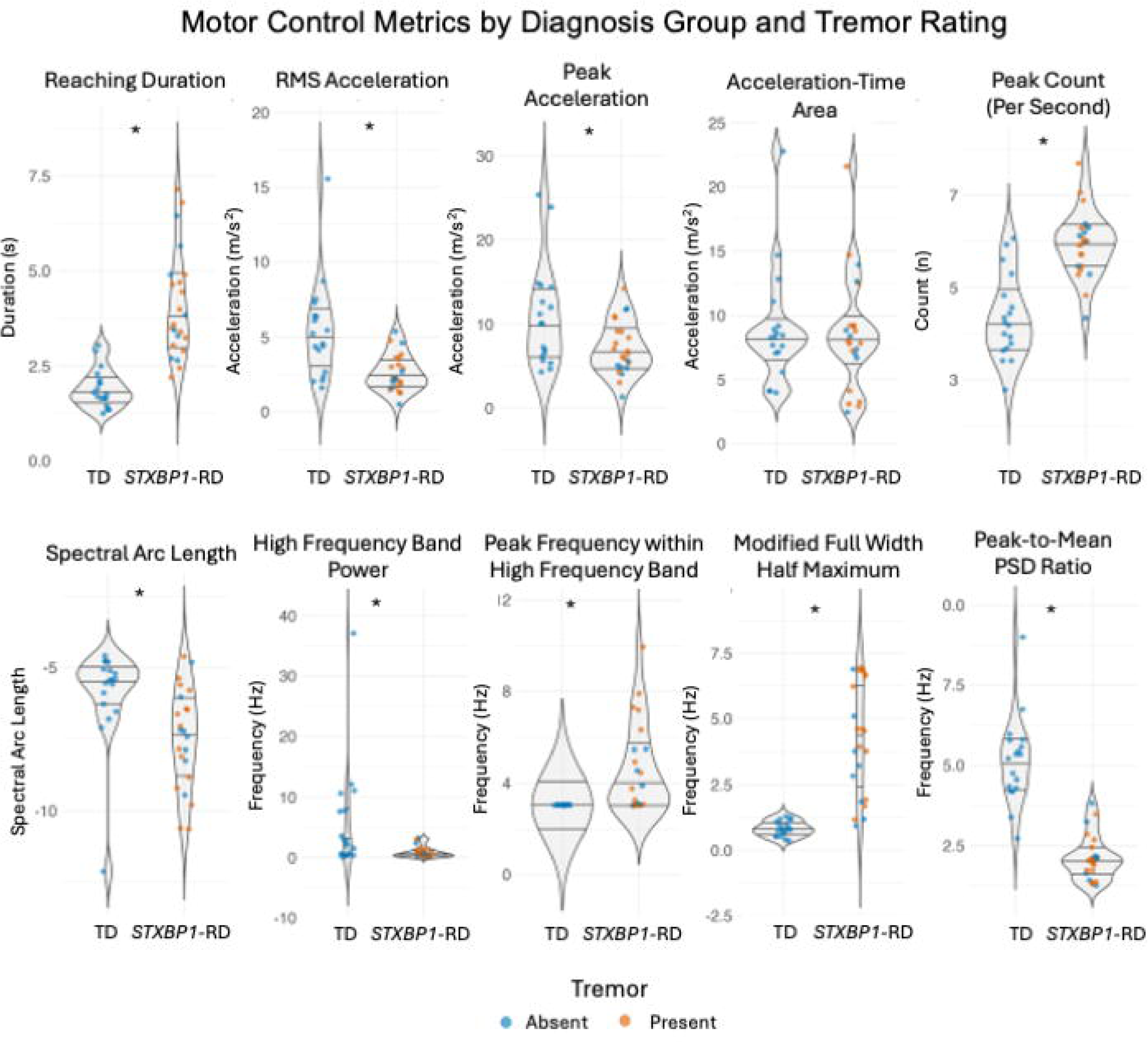
Motor Control Metrics Distinguished *STXBP1*-RD from TD Controls. Sensor metrics were evaluated from the faster side (i.e., dominant) forearm sensor. The tremor rating from the S-CSA is indicated as absent in blue or present in orange; tremor was absent in the TD group. * Indicates *P* <.05. Abbreviations: Typical Development = TD, *STXBP1*-Related Disorders = *STXBP1*-RD, Root Mean Squared = RMS, Power Spectral Density = PSD.

**Table 2.**
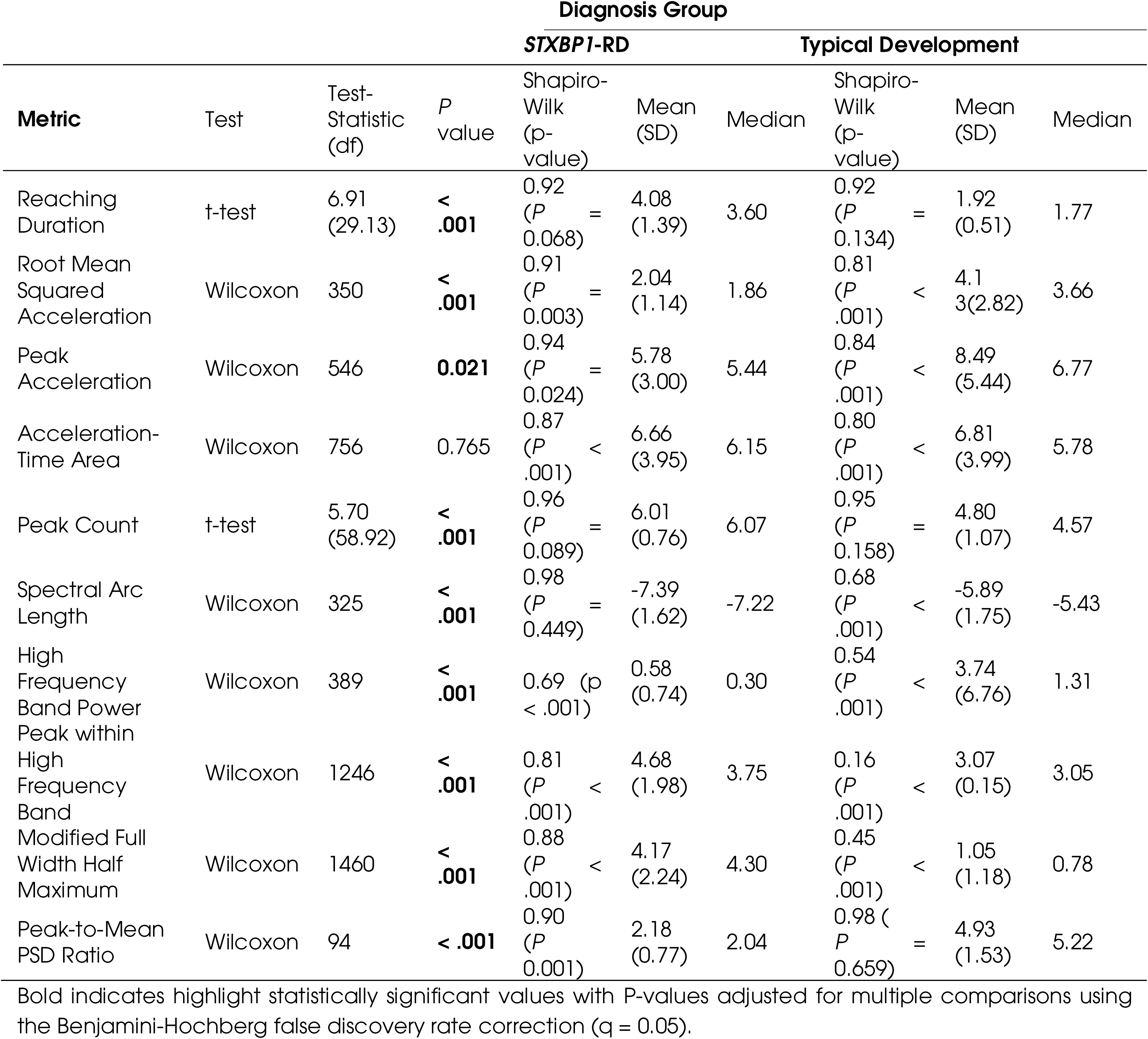
Sensor Derived Metrics Compared Between Diagnosis Groups.

Similarly, measures of smoothness were also different between groups. Peak count was significantly higher in participants with *STXBP1*-RD (t(58.92) = 5.70, *P* < .001; *STXBP1*-RD: M = 6.01, SD = 0.76; TD: M = 4.80, SD = 1.07). Participants with *STXBP1*-RD also exhibited significantly lower spectral arc length values, another measure of smoothness (V = 325, *P* < .001; *STXBP1*-RD: M = −7.39, SD = 1.62; TD: M = −5.89, SD = 1.75). Together, these smoothness-related findings indicate that individuals with *STXBP1*-RD demonstrate more fragmented and less smooth movement patterns during goal-directed reaching, consistent with impaired motor coordination.

Tremor was identified in 65% (15/23) of participants with *STXBP1*-RD who had evaluable sensor data during the clinical examination. The tremor metrics were selected to be robust to the absence of tremor, allowing inclusion of both participants with TD and *STXBP1*-RD without tremor; therefore, the entire cohort remained the focus of this analysis. Not surprisingly, tremor characteristics were significantly different between groups.

High Frequency Band Power was significantly higher for participants with TD compared to participants with *STXBP1*-RD (V = 389, *P* < .001; TD: M = 3.74, SD = 6.76), reflecting greater amplitude in the low frequency band, (i.e., voluntary goal directed movement) followed by a gradual decline across the high frequency band. In contrast, participants with *STXBP1*-RD showed a steeper decline in the low frequency band, consistent with lower high frequency band power (*STXBP1*-RD: M =0.58 SD = 0.74). The peak within the high frequency band for participants with TD occurred at the lower threshold of the band (TD: M = 3.07, SD = 0.15) and was significantly lower than for the *STXBP1*-RD group (V = 1245.5, *P* < .001; M =4.68, SD = 1.98) who often demonstrated oscillations (i.e., tremor) in this range. Participants with *STXBP1*-RD also exhibited broader FWHM_M_ (V = 1460, *P* < .001; *STXBP1*-RD: M =4.17, SD = 2.24; TD: M = 1.05, SD = 1.18) and a lower tremor peak-to-mean PSD ratio (V = 94, *P* < .001; *STXBP1*-RD: M =2.18 SD = 0.77 TD: M = 4.93, SD = 1.53). Taken together, these findings indicate that the tremor observed in the *STXBP1*-RD is an irregular signal consistent with a disorganized or unstable rhythmic movement.

Z-score analyses highlighted distinct differences between TD and *STXBP1*-RD groups across motor control metrics of duration, intensity, and smoothness, and metrics of tremor characteristics (**Figure 4**). Participants with TD were consistently clustered around the reference mean, while participants with *STXBP1*-RD showed broader distributions. For metrics of tremor-characteristics, participants with *STXBP1*-RD, particularly those with clinically identified tremor, demonstrated higher and more variable Z-scores, with FWHM_M_ and the tremor peak-to-mean PSD ratio showing the greatest group separation. At the individual level, most participants with *STXBP1*-RD had at least one metric beyond 2 SD of the TD reference, and several exceeded 3 SD. Deviations across movement constructs highlight the broad impact of *STXBP1*-RD on motor control.

**Figure 4.**
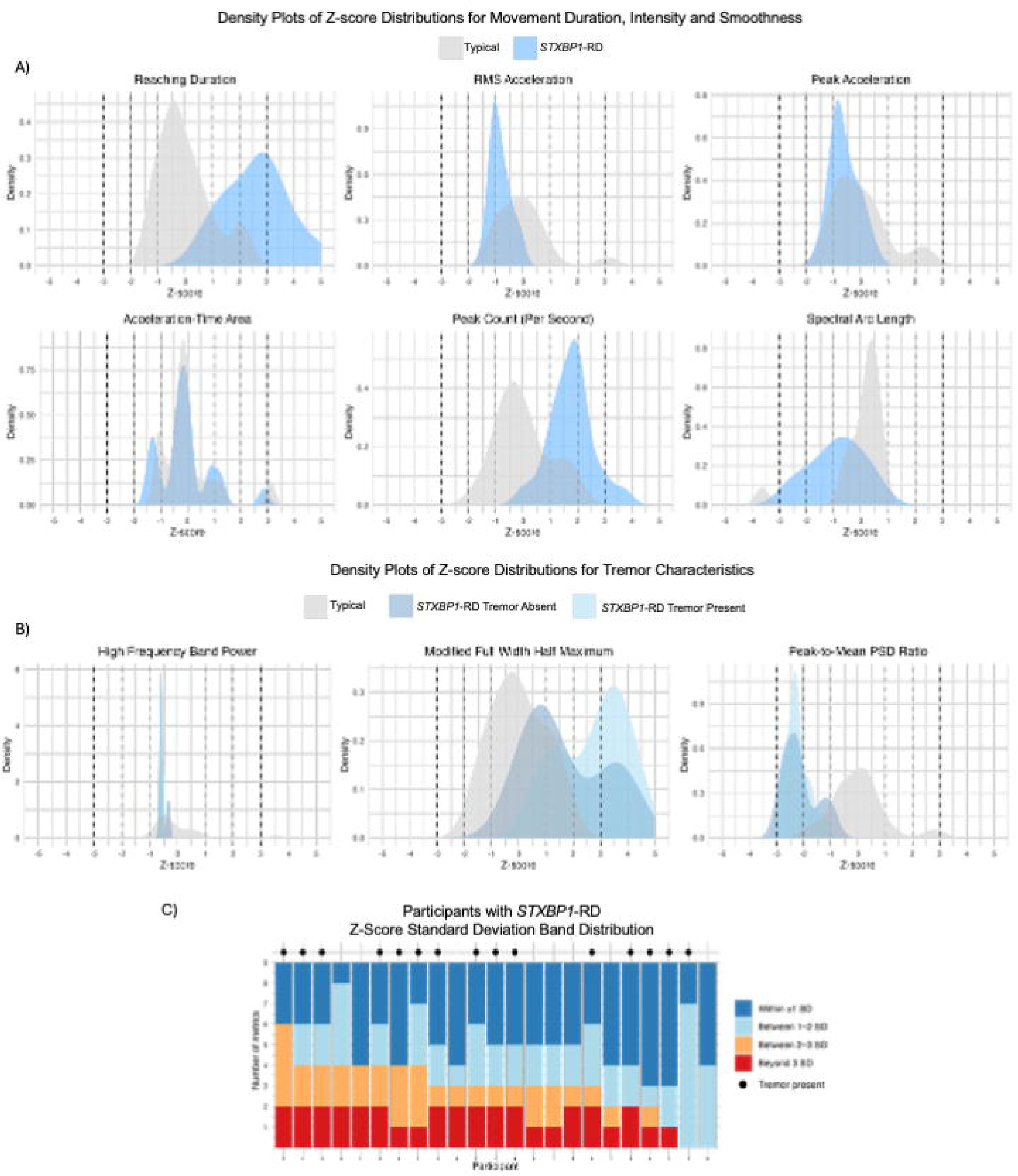
Z-Score Distributions of Motor Control Metrics. **(A**) Distribution of Z-scores for motor control metrics of duration, intensity and smoothness in participants with *STXBP1*-RD relative to the reference group with typical development (TD). For each metric, Z-scores were calculated using the mean and standard deviation of the TD group. Densities are shown for participants with TD (light gray) and participants with *STXBP1*-RD (steel blue). (B) For tremor characteristic metrics, participants with *STXBP1*-RD are further stratified by tremor presence (sky blue = Tremor Present, dark blue = Tremor Absent). Dashed vertical lines indicate ±1 SD (light gray), ±2 SD (dark gray), and ±3 SD (black) relative to the reference group with TD. The area under each density curve is normalized to 1, reflecting the relative distribution of participants across Z-scores. (**C**) Distribution of participants by Z-score deviation bands relative to the reference group with TD. Each bar represents the proportion of individuals with at least one metric falling within ±1 SD, 1–2 SD, 2–3 SD, or >3 SD. Most participants with *STXBP1*-RD had at least one metric beyond ±2 SD. Tremor presence was denoted with a black dot. Abbreviations: Typical Development = TD, *STXBP1*-Related Disorders = *STXBP1*-RD, Root Mean Squared = RMS, Power Spectral Density = PSD.

### Motor Control Metrics Were Predictive of Tremor Using Supervised Machine Learning

We then evaluated if the sensor-derived metrics of motor control could predict the presence of tremor, as measured by the S-CSA standardized clinical scale, using a supervised Random Forest machine learning approach. The TD control group was included with an absent tremor rating, as were participants with *STXBP1*-RD without clinically identified tremor. We tested multiple combinations of sensors, including forearm, upper arm, dominant and non-dominant limbs, to determine which configuration yielded the highest area under the curve (AUC) in Receiver Operating Characteristic (ROC) analyses. For each sensor configuration, we also identified the five most predictive sensor metrics. The AUC values ranged from 0.76 using a single sensor on the non-dominant forearm to 0.87 using all four sensors (bilateral forearm and upper arm sensors; **Figure 5A**). Across all sensor combinations, the features with the highest mean importance were FWHM_M_ (48.6 ± 37.5), reaching duration (39.9 ±32.9), peak-to-mean PSD ratio (37.3 ± 24.6), peak count (34.5 ± 33.8), and peak within high frequency power band (31.9 ± 34.2). These features consistently produced the largest drops in model accuracy when their values were permuted, indicating strong predictive value for distinguishing participants with and without tremor on the S-CSA (**Figure 5B**). In summary, motor control metrics were highly predictive of the presence of tremor with several features consistently emerging as predictors.

**Figure 5.**
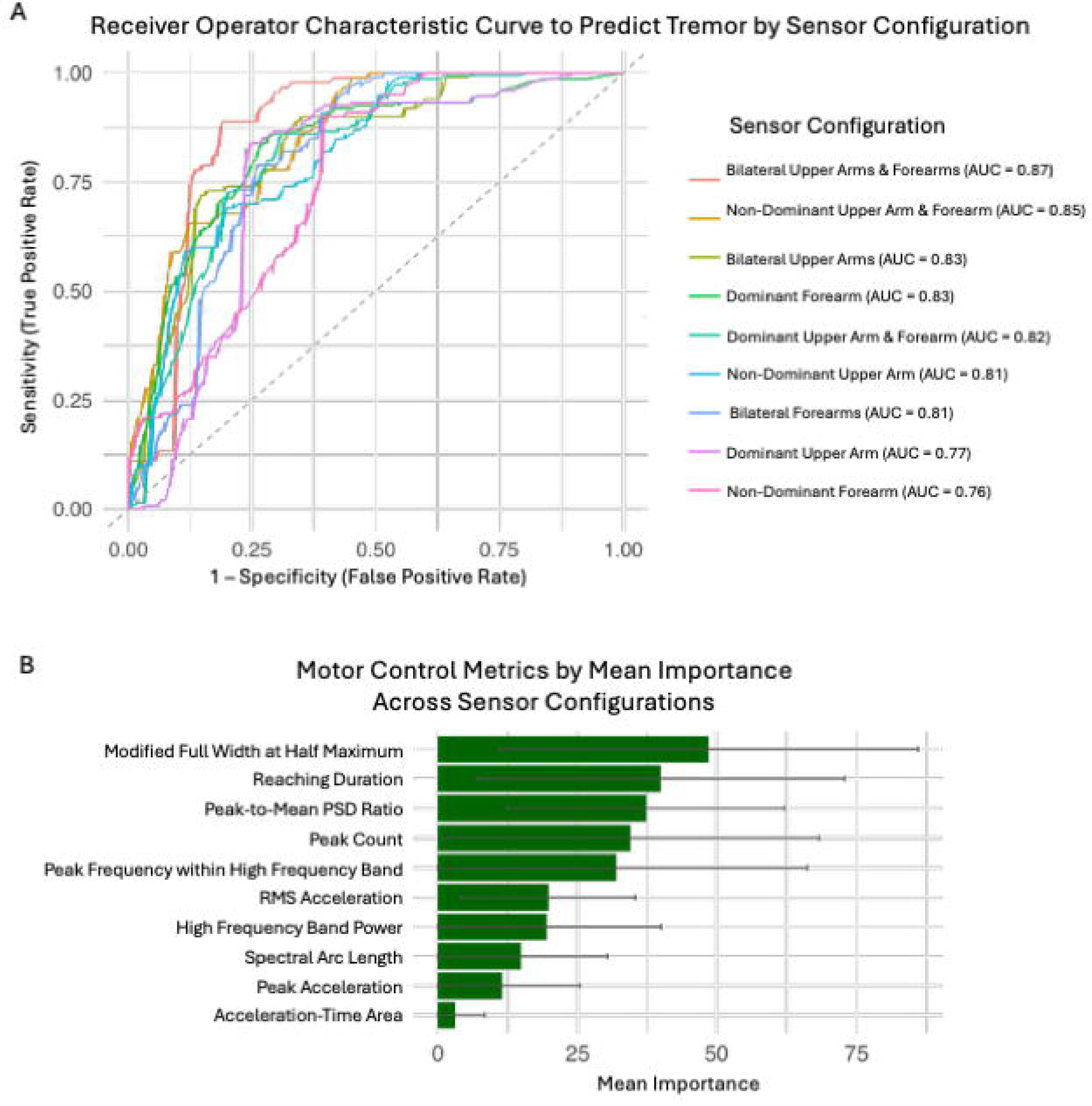
Supervised Machine Learning Predicted Tremor. **(A**) Receiver Operating Characteristic curves showing model performance predicting tremor for each combination of faster/slower reaching side and upper arm/ forearm sensors. (**B**) Mean importance of sensor-derived motor control metrics across all models, highlighting the features that most consistently contributed to tremor classification. Error bars represent ± standard deviation. Abbreviations: Area under the curve = AUC, Root Mean Squared = RMS, Power Spectral Density = PSD.

### A Subset of Sensor-Derived Motor Control Metrics Correlated with Developmental Outcomes

A subset of sensor-derived motor control metrics showed significant relations with standardized developmental assessments including the GMFM-66, and subscales of the Bayley-4 and PDMS-3 (**Figure 6**). Specifically, increased spectral arc length values (i.e., increased smoothness) were associated with higher scores across multiple developmental domains, including gross and fine motor skills (PDMS-3: Hand Manipulation: ρ = 0.464, *P* = 0.045; Eye-Hand Coordination: ρ = 0.489, *P* = 0.034; Bayley-4: Fine Motor: ρ = 0.558, *P* = 0.016; Gross Motor: ρ = 0.505, *P* = 0.033), and expressive communication (Bayley-4: Expressive Communication: ρ = 0.530, *P* = 0.024). Reaching duration (slower reaching) was negatively correlated with assessment scores across all domains (ρ range: –0.502 to –0.696; *P* range = 0.001–0.034), with the exception of receptive communication which did not reach statistical significance (ρ = –0.457, *P* = 0.056), suggesting that more efficient, faster reaching movements to a target was related to more advanced motor skills. Similarly, acceleration-time area (i.e., increased intensity) was positively related to fine motor scores (Bayley-4 Fine Motor: ρ = –0.560, *P* = 0.016), indicating that greater movement intensity during reaching was associated with more advanced fine motor skills.

**Figure 6.**
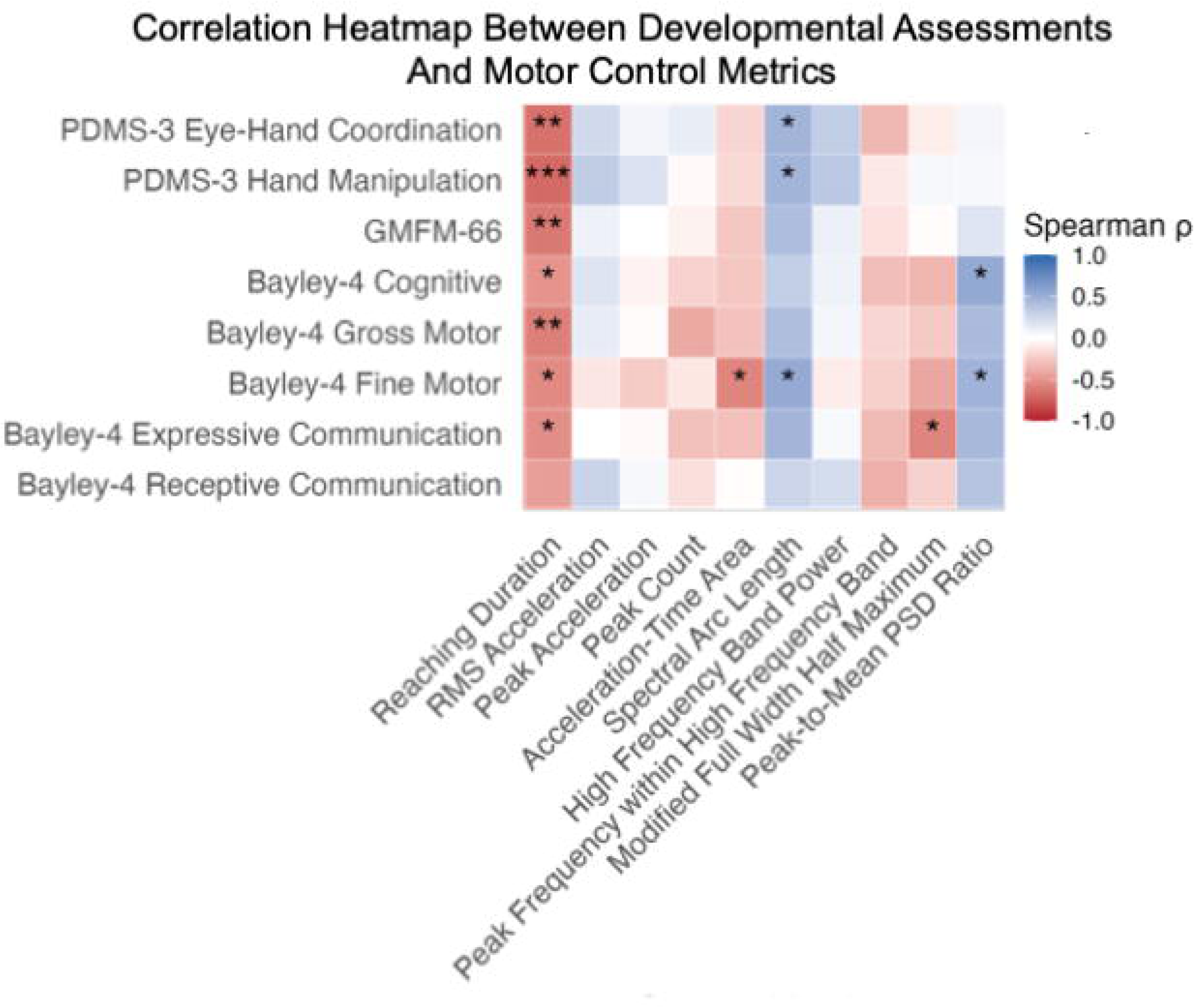
Spearman Correlation Coefficient between Developmental Assessments and Sensor-Derived Motor Control Metrics. Significant relations were observed between developmental assessment domains and motor control metrics. * Indicates *P* <.05, ** indicates *P* <.01, *** indicates *P* <.001. Abbreviations: Peabody Developmental Motor Scales, 3^rd^ Edition = PDMS-3, Gross Motor Function Measure, 66 items = GMFM-66; Bayley Scales of Infant and Toddler Development 4^th^ Edition = Bayley-4, Root Mean Squared = RMS, Power Spectral Density = PSD.

In addition to metrics of duration, intensity, and smoothness, several metrics of tremor characteristics also showed significant relations with developmental outcomes. Lower peak-to-mean PSD ratios, indicating less defined, more broadband tremor activity, were associated with poorer fine motor (ρ = 0.476, *P* = 0.046) and cognitive scores (ρ = 0.513, *P* = 0.029). Likewise, broader tremor peaks (i.e., higher FWHM_M_), indicative of less stable and more variable tremor patterns, were linked to lower fine motor (ρ = –0.473, *P* = 0.047) and expressive communication (ρ = –0.567, *P* = 0.014) outcomes. These findings suggest that more broadband, irregular frequency tremor is related to poorer developmental performance.

### Tremor was More Prevalent in Individuals with *STXBP1*-RD Compared to Other NDDs

Given the consistent reports of tremor in *STXBP1*-RD and the relations between Tremor Characteristics and developmental skill attainment, we also evaluated the prevalence of tremor in individuals with *STXBP1*-RD relative to other NDDs across different datasets. In a previously published large retrospective cohort study of 534 individuals with *STXBP1*-RD, tremor and ataxia were reported in 23.8% and 24.7% of individuals respectively, while abnormality of movement was present in 53.2% of individuals.^10^ Using the Citizen Health dataset, which aggregates real-world clinical information from electronic medical records (EMR) across DEEs of assumed genetic origin, abnormal movement was reported in 95.1% of individuals with *SCN8A*-RD, 73.9% with *SYNGAP1*-RD, and 97.8% with *STXBP1*-RD. In contrast, tremor was reported in 43.9% of *SCN8A*-RD, 14.5% of *SYNGAP1*-RD, and 66.3% of *STXBP1*-RD, while ataxia was reported in 41.5%, 18.1%, and 47.8%, respectively. Thus, although tremor is generally uncommon in DEEs, it occurs up to five times more frequently in *STXBP1*-RD. In the STARR natural history data, 68.5% (50/73) of individuals evaluated have been observed to have tremor during at least one study visit using the S-CSA. When evaluated in the context of a natural history study, the rates of tremor were similar or higher than the rates reported through retrospective datasets. Of note, in clinical experience, families of individuals with *STXBP1*-RD report unique features associated with tremor, including that tremor is most often observed upon waking or in the setting of a fever or illness. Of note, in the STARR natural history study cohort, there were 5 participants with instances of concern for a seizure that the provider noted was more likely to be tremor. These findings highlight the prevalence and unique presentation of tremor in *STXBP1*-RD, motivating our sensor-based analyses to quantify motor control and specifically tremor characteristics and their relation to developmental outcomes.

## Discussion

Here, we evaluated neuromotor control using wearable sensors during a structured reaching task for individuals with *STXBP1*-RD. The work demonstrates a novel approach to objectively quantify motor control, including highly prevalent tremor characteristics, in this population. We found that measures of duration, intensity, smoothness and tremor characteristics not only distinguished between individuals with STXBP1-RD and their peers with TD, but predicted the presence of tremor and were also associated with functional performance across several developmental domains. This work lays the foundation for future research that can expand to other settings, such as non-clinical naturalistic environments and supports the development of objective biomarkers to facilitate clinical trial readiness in *STXBP1*-RD.

While wearable sensors have been used to assess tremor, ataxia, and other movement disorders in individuals with various neurological disorders,^22,26,45^ these studies predominantly relied on complex tasks, such as drawing an Archimedes spiral, finger-to-nose tasks, or different combinations of finger tapping.^46–49^ These testing paradigms are not feasible for children who are not able to follow multi-step directions, including children with *STXBP1*-RD. We addressed this problem by recording natural movement during a simple, but standardized, reaching task and calculating precise measures of motor control. The reaching paradigm replicates usual motor behavior, the wearable sensors are inobtrusive and the approach **can be implemented** within or outside of a clinical environment.

Our study has four main findings. First, we showed that neuromotor control in the *STXBP1-RD* population can be quantified using wearable sensors. The structured reaching task **was feasible for most participants (83.9%), who completed** at least three voluntary reaches for objects with at least one arm. Motor control metrics distinguished participants with *STXBP1*-RD and TD groups during the simple reaching task. The reaching movement frequency characteristics revealed a broadband spectrum, with power distributed across multiple frequencies and no single dominant peak, as well as irregular burst activity. These patterns are indicative of disorganized or unstable rhythmic movements, consistent with features of rhythmic myoclonus. This aligns with the findings of Loussouarn et al.^18^, who conducted upper extremity neurophysiological evaluations using electromyography and observed irregular electromyography burst durations that followed a somewhat rhythmic pattern. This upper limb muscle activity typically occurred within the 4–10 Hz frequency range and the authors therefore characterized the *STXBP1*-RD tremor as a pseudo-rhythmic subcortical myoclonus. Furthermore, the investigators noted a predominance of tremor in distal or proximo-distal anatomical limb regions. Consistent with this evidence, our machine learning models demonstrated greater predictive accuracy for tremor using forearm sensor data compared to upper arm data alone, suggesting increased tremor expression distally. Collectively, these findings corroborate previous neurophysiological characterizations and support the implementation of unobtrusive, easy-to-collect wearable sensor-based assessments to quantify movement disorders in individuals with *STXBP1*-RD.

Second, we demonstrated that sensor-derived motor control metrics distinguished individuals with *STXBP1*-RD from those with TD. This suggests that there are measurable differences in motor control between groups that are not entirely attributed to the presence of tremor. We observed significant differences in reaching duration, intensity, and smoothness, in addition to differences in tremor characteristics. These findings indicate that *STXBP1*-RD may impact multiple constructs of motor control and coordination, a finding which has also been described in individuals with ASD.^45,50,51^ Decreased smoothness (i.e., spectral arc length), for example, may be a manifestation of impairments in motor sequencing, temporal consistency or target path-deviations. Based on these findings, further exploration of the breadth and depth of motor coordination impairments in *STXBP1*-RD is warranted.

Third, we also showed that motor control metrics are meaningfully associated with developmental outcomes in *STXBP1*-RD. Importantly, the associations spanned multiple domains, linking smoother, faster reaching movements and greater movement intensity with better performance on standardized assessments of gross motor, fine motor, cognitive, and language skills. The association between tremor frequency characteristics and developmental skills further clarifies the functional impact of upper limb tremor within this population. Families frequently report daily fluctuations in their child’s motor coordination, including variability in tremor, which can be difficult to capture with clinical assessments. Future work should investigate whether changes in sensor-derived motor control metrics are consistent with this caregiver-reported variability, as this would strengthen their utility. This work demonstrates the potential of **sensor-derived** motor control **metrics to provide sensitive, objective, and functionally relevant endpoints that extend beyond conventional clinical scales.**

Fourth, we characterized tremor and additional movement disorders in individuals with *STXBP1*-RD. While movement disorders are commonly described in NDDs with assumed genetic origin, tremor is not observed consistently across various diagnoses. Importantly, tremor in the *STXBP1*-RD population is common and present in up to 68.5% of individuals in prospective natural history studies yet significantly underreported in retrospective datasets. These results highlight the importance of objective measures to classify movement disorders beyond phenotypic descriptions.

Given that our aim was to objectively quantify movement disorders in individuals with significant developmental impairments, our study has several limitations. We selected three reaching intervals per participant in order to optimally balance feasibility and data reliability. This meant the total amount of evaluable data from each participant was relatively small and constrained our ability to compute certain sensor-derived metrics that require a longer time series (more than 200 data points) for reliable calculation. Specifically, nonlinear measures of signal complexity and variability such as sample entropy (reflecting signal predictability), spectral entropy (representing frequency dispersion), and fractal dimension (a quantification of self-similarity and irregularity in movement patterns), could not be reliably calculated. Future studies should consider incorporating longer recording durations of natural, unstructured movement behavior to minimize participant exclusion due to significant cognitive or physical impairments and to increase the duration of analyzable data. Additionally, wearable sensors with higher sampling rates should be explored to support robust estimation of metrics requiring more data points. Finally, the present study does not explore sensor-derived metrics longitudinally, and future work is needed to evaluate their consistency across time and their response to intervention. Anecdotally, some families have reported that their child’s tremor has become more pronounced over time; further studies evaluating changes across individuals and across time are needed to explore this question.

In summary, our findings show that wearable sensors paired with a simple reaching task can accurately capture neuromotor control and tremor, distinguish individuals with STX and TD, correlate with developmental abilities, and reveal that tremor in *STXBP1*-RD is prevalent yet often overlooked, emphasizing the importance of objective evaluation. This study addresses the critical need for objective, scalable outcome measures to support clinical trial readiness for individuals with *STXBP1*-RD. Importantly, wearable sensors also hold the potential for remote data collection, allowing for the recording of motor control in natural environments and enabling the inclusion of individuals who are often unable to participate in natural history studies and clinical trials due to travel barriers or to significant cognitive or motor impairments. Future work should focus on expanding assessments to include more naturalistic, unstructured tasks, optimizing sensor placement and sampling parameters, and validating remote protocols to ensure broader inclusion and the opportunity for longitudinal monitoring.

## Data availability

The data that support the findings of this study are available from the corresponding author, upon reasonable request.

## Acknowledgements

The authors would like to acknowledge the individuals and their families who took part in this study as well as the *STXBP1* Foundation.

## Supplementary material

Supplementary material is available at *Brain* online.

## References

1. Oliver KL, Scheffer IE, Bennett MF, Grinton BE, Bahlo M, Berkovic SF. Genes4Epilepsy: An epilepsy gene resource. Epilepsia. 2023-05-01 2023;64(5):1368–1375. doi:10.1111/epi.17547

2. JL M, JH M, J X, et al. Clinical signatures of SYNGAP1-related disorders through data integration - PubMed. Genetics in medicine : official journal of the American College of Medical Genetics. 2025 Jun;27(6) doi:10.1016/j.gim.2025.101419

3. Magielski JH, Cohen S, Kaufman MC, et al. Deciphering the Natural History of SCN8A-Related Disorders. Neurology. May 13 2025;104(9):e213533. doi:10.1212/wnl.0000000000213533

4. Papandreou A, Danti FR, Spaull R, Leuzzi V, Mctague A, Kurian MA. The expanding spectrum of movement disorders in genetic epilepsies. Developmental Medicine & Child Neurology. 2020-02-01 2020;62(2):178–191. doi:10.1111/dmcn.14407

5. Yuan M, Wang X, Yang Z, Luo H, Gan J, Luo R. Advances in genetic developmental and epileptic encephalopathies with movement disorders. Acta Epileptologica. 2025-12-01 2025;7(1) doi:10.1186/s42494-024-00194-z

6. Van Der Veen S, Tse GTW, Ferretti A, et al. Movement Disorders in Patients With Genetic Developmental and Epileptic Encephalopathies. Neurology. 2023-11-07 2023;101(19):e1884–e1892. doi:10.1212/wnl.0000000000207808

7. N M, MJ B, BL P. Accelerating therapeutic development and clinical trial readiness for STXBP1 and SYNGAP1 disorders - PubMed. Current problems in pediatric and adolescent health care. 2024 Aug;54(8) doi:10.1016/j.cppeds.2024.101576

8. Goss JR, Prosser B, Helbig I, Rigby CS. STXBP1: fast-forward to a brighter future – a patient organization perspective. Therapeutic Advances in Rare Disease. 2024 Jun 18;5doi:10.1177/26330040241257221

9. Saitsu H, Kato M, Mizuguchi T, et al. De novo mutations in the gene encoding STXBP1 (MUNC18-1) cause early infantile epileptic encephalopathy. NATURE GENETICS. 2008-06-01 2008;40(6):782–788. doi:10.1038/ng.150

10. Xian J, Parthasarathy S, Ruggiero SM, et al. Assessing the landscape of STXBP1-related disorders in 534 individuals. Brain. 2021 Nov 23;145(5) doi:10.1093/brain/awab327

11. Sullivan KR, Ruggiero SM, Xian J, et al. A disease concept model for STXBP1Lrelated disorders. Epilepsia Open. 2023 Apr 27;8(2) doi:10.1002/epi4.12688

12. Kim YO, Korff CM, Villaluz MMG, et al. Head stereotypies in STXBP1 encephalopathy. Developmental Medicine & Child Neurology. 2013/08/01;55(8) doi:10.1111/dmcn.12197

13. E K, E K, C O, et al. Motor phenotyping in a Greek cohort of patients with neonatal and infantile onset developmental and epileptic encephalopathy - PubMed. European journal of paediatric neurology : EJPN : official journal of the European Paediatric Neurology Society. 2025 Mar;55doi:10.1016/j.ejpn.2025.03.001

14. Deprez L, Weckhuysen S, Holmgren P, et al. Clinical spectrum of early-onset epileptic encephalopathies associated with STXBP1 mutations. Neurology. Sep 28 2010;75(13):1159–65. doi:10.1212/WNL.0b013e3181f4d7bf

15. Di Meglio C, Lesca G, Villeneuve N, et al. Epileptic patients with de novo STXBP1 mutations: Key clinical features based on 24 cases. Epilepsia. Dec 2015;56(12):1931–40. doi:10.1111/epi.13214

16. Mignot C, Moutard ML, Trouillard O, et al. STXBP1-related encephalopathy presenting as infantile spasms and generalized tremor in three patients. Epilepsia. Oct 2011;52(10):1820–7. doi:10.1111/j.1528-1167.2011.03163.x

17. Keogh M, Daud D, Pyle A, et al. A novel de novo STXBP1 mutation causes mitochondrial complex I deficiency and late onset juvenile onset parkinsonism. Neurogenetics. 2014 Nov 25;16(1) doi:10.1007/s10048-014-0431-z

18. Loussouarn A, Doummar D, Beaugendre Y, et al. Tremor-like subcortical myoclonus in STXBP1 encephalopathy. Eur J Paediatr Neurol. Sep 2021;34:62–66. doi:10.1016/j.ejpn.2021.06.005

19. Elble RJ, Ondo W. Tremor rating scales and laboratory tools for assessing tremor. Journal of the Neurological Sciences. 2022/04/15/ 2022;435:120202. 10.1016/j.jns.2022.120202

20. Saute JA, Donis KC, Serrano-Munuera C, et al. Ataxia rating scales--psychometric profiles, natural history and their application in clinical trials. Cerebellum. Jun 2012;11(2):488–504. doi:10.1007/s12311-011-0316-8

21. Schouwstra KJ, Polet SS, Hbrahimgel S, et al. Application of the Scale for Assessment and Rating of Ataxia in toddlers. European Journal of Paediatric Neurology. 2022/09/01;40doi:10.1016/j.ejpn.2022.07.001

22. Lobo MA, Hall ML, Greenspan B, Rohloff P, Prosser LA, Smith BA. Wearables for Pediatric Rehabilitation: How to Optimally Design and Use Products to Meet the Needs of Users. Phys Ther. Jun 1 2019;99(6):647–657. doi:10.1093/ptj/pzz024

23. Locatelli P, Alimonti D, Traversi G, Re V. Classification of Essential Tremor and Parkinson’s Tremor Based on a Low-Power Wearable Device. Electronics. 2020-10-15 2020;9(10):1695. doi:10.3390/electronics9101695

24. Li J, Zhu H, Li J, et al. A Wearable Multi-Segment Upper Limb Tremor Assessment System for Differential Diagnosis of Parkinson’s Disease Versus Essential Tremor. IEEE Transactions on Neural Systems and Rehabilitation Engineering. 2023-01-01 2023;31:3397–3406. doi:10.1109/tnsre.2023.3306203

25. Mishra RK, Nunes AS, Enriquez A, et al. At-home wearable-based monitoring predicts clinical measures and biological biomarkers of disease severity in Friedreich’s Ataxia. Communications Medicine. 2024-10-29 2024;4(1) doi:10.1038/s43856-024-00653-1

26. Knudson KC, Gupta AS. Assessing Cerebellar Disorders with Wearable Inertial Sensor Data Using Time-Frequency and Autoregressive Hidden Markov Model Approaches. Sensors. 2022;22(23):9454. doi:10.3390/s22239454

27. Amiri A, Peltier N, Goldberg C, et al. WearSense: Detecting Autism Stereotypic Behaviors through Smartwatches. Healthcare. 2017-02-28 2017;5(1):11. doi:10.3390/healthcare5010011

28. Using a Smartwatch to Detect Stereotyped Movements in Children With Developmental Disabilities. IEEE Access. 2017;5 doi:10.1109/ACCESS.2017.2689067

29. Vescio B, Quattrone A, Nisticò R, Crasà M, Quattrone A. Wearable Devices for Assessment of Tremor. Front Neurol. 2021;12:680011. doi:10.3389/fneur.2021.680011

30. van den Brandhof EL, Tuitert I, Madelein van der Stouwe AM, et al. Explainable machine learning for movement disorders - Classification of tremor and myoclonus. Comput Biol Med. Jun 2025;192(Pt B):110180. doi:10.1016/j.compbiomed.2025.110180

31. Prosser LA, Skorup J, Pierce SR, et al. Locomotor learning in infants at high risk for cerebral palsy: A study protocol. Frontiers in Pediatrics. 2023;11doi:10.3389/fped.2023.891633

32. Ricotti V, Kadirvelu B, Selby V, et al. Wearable full-body motion tracking of activities of daily living predicts disease trajectory in Duchenne muscular dystrophy. Nature Medicine. 2023;29(1):95–103. doi:10.1038/s41591-022-02045-1

33. Shirai S, Yabe I, Takahashi-Iwata I, et al. The Responsiveness of Triaxial Accelerometer Measurement of Gait Ataxia Is Higher than That of the Scale for the Assessment and Rating of Ataxia in the Early Stages of Spinocerebellar Degeneration. The Cerebellum. 2019;18(4):721–730. doi:10.1007/s12311-019-01025-5

34. Datavyu Team. Datavyu: A Video Coding Tool. New York University: Databrary Project; 2014.

35. Angione K, Abbott M, Stringfellow M, et al. Performance of Clinical Severity Assessment Measures on Several Neurogenetic Populations. www.aesnet.org; 2023:

36. Abbott M, Angione K, Stringfellow M, et al. Cortical Visual Impairment Across a Range of Neurodevelopmental Disorders: Clinical Characterization, Diagnostic Tool Evaluation, and Association with Developmental Outcomes. J Child Neurol. Aug 6 2025:8830738251361698. doi:10.1177/08830738251361698

37. Demarest S, Pestana-Knight EM, Olson HE, et al. Severity Assessment in CDKL5 Deficiency Disorder. Pediatr Neurol. Aug 2019;97:38–42. doi:10.1016/j.pediatrneurol.2019.03.017

38. Bayley N, Aylward GP. Bayley Scales of Infant and Toddler Development. fourth ed: Pearson; 2019.

39. Russell DJ, Rosenbaum PL, Wright M, Avery LM. Gross Motor Function Measure (GMFM-66 and GMFM-88) User’s Manual. 2 ed: Mac Keith Press; 2013.

40. Folio MR, Fewell RR. Peabody Developmental Motor Scales. vol 3rd Edition. Pro-Ed; 2023.

41. Health C. https://www.citizen.health

42. Robinson PN, Köhler S, Bauer S, Seelow D, Horn D, Mundlos S. The Human Phenotype Ontology: a tool for annotating and analyzing human hereditary disease. Am J Hum Genet. Nov 2008;83(5):610–5. doi:10.1016/j.ajhg.2008.09.017

43. Pierce S, Cunningham K, Coyne J, et al. Validating STXBP1 & SYNGAP1 Related Disorders: Assessing the Feasibility of Developmental Outcome Measures. 2024:

44. Rosenbaum PL, Walter SD, Hanna SE, et al. Prognosis or gross motor function in cerebral palsy - Creation of motor development curves. Jama-Journal of the American Medical Association. Sep 2002;288(11):1357–1363. doi:10.1001/jama.288.11.1357

45. Wilson RB, Vangala S, Elashoff D, Safari T, Smith BA. Using Wearable Sensor Technology to Measure Motion Complexity in Infants at High Familial Risk for Autism Spectrum Disorder. Sensors. 2021-01-17 2021;21(2):616. doi:10.3390/s21020616

46. Ali SM, Arjunan SP, Peter J, et al. Wearable Accelerometer and Gyroscope Sensors for Estimating the Severity of Essential Tremor. IEEE J Transl Eng Health Med. 2024;12:194–203. doi:10.1109/jtehm.2023.3329344

47. Lukšys D, Jonaitis G, Griškevičius J. Quantitative Analysis of Parkinsonian Tremor in a Clinical Setting Using Inertial Measurement Units. Parkinsons Dis. 2018;2018:1683831. doi:10.1155/2018/1683831

48. van der Linden C, Berger T, Brandt GA, et al. Accelerometric Classification of Resting and Postural Tremor Amplitude. Sensors (Basel). Oct 21 2023;23(20) doi:10.3390/s23208621

49. Paredes-Acuna N, Utpadel-Fischler D, Ding K, Thakor NV, Cheng G. Upper limb intention tremor assessment: opportunities and challenges in wearable technology. J Neuroeng Rehabil. Jan 13 2024;21(1):8. doi:10.1186/s12984-023-01302-9

50. Fournier KA, Hass CJ, Naik SK, et al. Motor Coordination in Autism Spectrum Disorders: A Synthesis and Meta-Analysis. Journal of Autism and Developmental Disorders 2010 40:10. 2010-03-02;40(10) doi:10.1007/s10803-010-0981-3

51. Wilson RB, Vangala S, Reetzke R, Piergies A, Ozonoff S, Miller M. Objective measurement of movement variability using wearable sensors predicts <scp>ASD</scp> outcomes in infants at high likelihood for ASD and ADHD. Autism Research. 2024-06-01 2024;17(6):1094–1105. doi:10.1002/aur.3150

