## Supplemental materials for "Quantification of neuromotor control in *STXBP1*-Related Disorders with wearable sensors"

### A. Bayley Scales of Infant and Toddler Development Age-Equivalent Scores

| Domain | Mean (SD) |
|---|---|
| **Bayley-4** | |
| Cognitive | 11.9 (7.4) |
| Gross Motor | 14.1 (8.1) |
| Fine Motor | 13.3 (8.0) |
| Expressive Communication | 11.7 (9.4) |
| Receptive Communication | 12.3 (8.3) |
| **PDMS-3** | |
| Eye-Hand Coordination | 9.2 (5.2) |
| Hand Manipulation | 9.9 (6.3) |
| **GMFM-66** | |
| GMFM-66 | 53.6 (11.0) |

Values are presented as mean (standard deviation). Bayley-4 and PDMS-3 scores represent age-equivalent scores in months for each domain. GMFM-66 represents the total score (range 0–100) for gross motor function, a typically developing 5-year-old would be expected to score 100.[44]
Abbreviations: GMFM-66, Gross Motor Function Measure-66; PDMS-3, Peabody Developmental Motor Scales, Third Edition; Bayley-4, Bayley Scales of Infant and Toddler Development, Fourth Edition.

B. Tremor Related Items on the *STXBP1*- Clinical Severity Assessment (S-CSA)

**Distribution:** 0= absent, 1 (or 33.333) = focal, one limb of the body, 2 (or 66.666) = segmental, affecting an entire side of the body or trunk/core, 3 (or 100) = generalized, affecting most of the body

**Duration:** 0 = absent, 1 (or 33.333) = intermittent, less than half of exam, 2 (or 66.666) = common, half or more than half of exam, 3 (or 100) = persistent, all of exam

**Functional impact:** 0 = absent, 1 (or 33.333) = Present but little to no interference with patient's fine or gross motor abilities, 2 (or 66.666) = Present and clearly is a contributor to patient's fine or gross motor disabilities, 3 (or 100) = Present and is the major or primary driver of patient's fine or gross motor disabilities

| C. Z-Scores by Participant | | | | | | | | | | |
|---|---|---|---|---|---|---|---|---|---|---|
| Diagnosis | Tremor Present | Reaching Duration | RMS Acceleration | Peak Acceleration | Acceleration-Time-Area | Peak Count | Spectral Arc Length | High Frequency Band Power | Modified Full Width Half Maximum | Peak to Mean PSD Ratio |
| STXBP1 | TRUE | 4.08 | -0.96 | -0.81 | -0.24 | 1.83 | -1.28 | -0.6 | 10.66 | -2.28 |
| STXBP1 | FALSE | 3.79 | -0.99 | -1 | -0.34 | 1.8 | 0.64 | -0.59 | 20.94 | -2.85 |
| STXBP1 | TRUE | 4.96 | -0.47 | -0.01 | 1.3 | 1.9 | -1.7 | -0.54 | 20.4 | -2.36 |
| STXBP1 | TRUE | 10.25 | -1.13 | -0.89 | 0.06 | 1.23 | -2.27 | -0.62 | 20.94 | -2.35 |
| STXBP1 | TRUE | 5.84 | -0.75 | -0.78 | 0.8 | 1.51 | -1.13 | -0.55 | 10.11 | -2.21 |
| STXBP1 | FALSE | 5.84 | -1.03 | -0.86 | -0.12 | 1.96 | -0.74 | -0.61 | 0.38 | -2.22 |
| STXBP1 | TRUE | 9.56 | -0.53 | -0.29 | 2.85 | 2.19 | -2.74 | -0.47 | 13.09 | -2.22 |
| STXBP1 | TRUE | 3.1 | -1.07 | -0.28 | -1.08 | 2.14 | 0.76 | -0.52 | 20.12 | -2.81 |
| STXBP1 | FALSE | 7.31 | -1.49 | -1.56 | -1.45 | 2.04 | -1.15 | -0.64 | 8.22 | -2.17 |
| STXBP1 | TRUE | 3.3 | -0.54 | 0.01 | -0.37 | 1.03 | -0.42 | -0.55 | 3.89 | -1.67 |
| STXBP1 | TRUE | 5.45 | -0.95 | -0.51 | -0.03 | 1.78 | -1.92 | -0.59 | 20.94 | -2.27 |
| STXBP1 | FALSE | 2.71 | -0.23 | 0.16 | 0.84 | 1.23 | -1.37 | -0.37 | 10.11 | -2.56 |
| STXBP1 | FALSE | 3 | -0.95 | -0.99 | -0.43 | 2.24 | -0.88 | -0.6 | 6.87 | -2.2 |
| STXBP1 | FALSE | 1.53 | 0 | 0.14 | 1.12 | 2.16 | -0.07 | -0.28 | 14.71 | -2.73 |
| STXBP1 | TRUE | 0.55 | -0.19 | 0.54 | 0.06 | 1.73 | -0.32 | -0.3 | 20.94 | -2.76 |
| STXBP1 | TRUE | 2.61 | -1.19 | -0.67 | -1.27 | 3.7 | -0.33 | -0.59 | 12.82 | -2.5 |
| STXBP1 | TRUE | 2.9 | -0.68 | -0.72 | 0.06 | 3 | -0.67 | -0.53 | 18.77 | -2.78 |
| STXBP1 | TRUE | 1.93 | -0.72 | -0.41 | -0.25 | 0.54 | 0.31 | -0.48 | 3.08 | -1.8 |
| STXBP1 | FALSE | 8.88 | -1.15 | -1.11 | -0.02 | 0 | -2.06 | -0.63 | 3.62 | -0.97 |
| STXBP1 | TRUE | 1.04 | -1.24 | -1.08 | -1.35 | 1.51 | 0.18 | -0.62 | 1.19 | -1.21 |
| STXBP1 | TRUE | 2.22 | -1.26 | -1.28 | -1.31 | 2.81 | 0.07 | -0.63 | 21.21 | -2.5 |
| STXBP1 | FALSE | 1.44 | -0.82 | -0.95 | -0.5 | 1.04 | -0.76 | -0.57 | 1.19 | -1.39 |
| STXBP1 | TRUE | 5.35 | -1.03 | -0.62 | -0.2 | 1.14 | -2.76 | -0.62 | 13.09 | -1.97 |
| TD | FALSE | -0.91 | 3.1 | 2.37 | 3.11 | 0.54 | 0.5 | 3.55 | -0.17 | 0.47 |
| TD | FALSE | -0.03 | 0.65 | 2.13 | 0.88 | -0.73 | -0.7 | 0.61 | 1.19 | -1.77 |

| | | | | | | | | | | |
|---|---|---|---|---|---|---|---|---|---|---|
| TD | FALSE | -0.42 | -0.04 | -0.14 | -0.14 | 1.05 | 0.31 | -0.24 | -0.17 | 0.31 |
| TD | FALSE | -0.52 | 0.02 | 0.05 | -0.06 | -0.78 | -0.37 | -0.35 | -0.44 | 0.14 |
| TD | FALSE | -0.52 | 1.02 | 0.65 | 1.29 | 1.9 | 0.65 | 0.24 | -0.98 | 0.15 |
| TD | FALSE | 0.75 | -1 | -0.91 | -1.08 | 0.01 | 0.41 | -0.58 | 0.11 | -0.43 |
| TD | FALSE | 2.22 | -1.15 | -1.02 | -1.11 | -0.45 | -3.63 | -0.63 | 1.73 | -0.58 |
| TD | FALSE | -1.31 | 0.58 | 0.57 | 0 | -0.38 | 0.66 | 0.73 | 1.19 | -0.69 |
| TD | FALSE | -1.11 | 0.22 | 0.61 | -0.27 | 1.75 | -0.52 | 0.22 | 0.38 | -0.62 |
| TD | FALSE | -1.11 | 0.3 | 0.18 | -0.1 | 0.11 | 0.47 | 0.56 | 0.92 | -0.7 |
| TD | FALSE | -0.52 | -0.31 | -0.64 | -0.4 | -0.11 | 0.27 | -0.47 | -0.98 | 0.46 |
| TD | FALSE | -0.33 | 0.32 | 0.28 | 0.48 | 0.25 | 0.22 | -0.31 | -0.98 | 1.17 |
| TD | FALSE | 0.26 | -0.3 | -0.7 | 0.06 | -1.02 | -0.21 | -0.57 | -1.52 | 2.82 |
| TD | FALSE | 0.26 | -0.38 | -0.59 | -0.12 | -1.02 | 0.02 | -0.55 | -1.52 | -0.29 |
| TD | FALSE | 1.93 | -0.83 | -0.78 | -0.41 | -0.61 | 0.68 | -0.61 | 0.38 | 0.2 |
| TD | FALSE | 1.14 | -0.93 | -0.85 | -0.75 | 1.39 | 0.28 | -0.59 | 1.46 | -1.28 |
| TD | FALSE | 0.46 | -1.02 | -1.08 | -1.08 | -0.17 | 0.78 | -0.6 | -0.17 | 0.05 |
| TD | FALSE | -0.23 | -0.25 | -0.13 | -0.29 | -1.73 | 0.19 | -0.42 | -0.44 | 0.6 |
